# Peer Support in Online Discussions of Male Infertility: A Natural Language Processing Study of Reddit

**DOI:** 10.64898/2026.03.09.26347980

**Authors:** Masuma Khatun, Naisarg Patel, Marina Loid, Aspasia Destouni, Prakash Lingasamy, Sajitha Lulu S, Maire Peters, Rajesh Sharma, Andres Salumets, Vijayachitra Modhukur

## Abstract

Infertility generates profound psychological and social distress for both women and men, yet men’s communicative experiences remain comparatively underexamined. Male infertility (MI) is often shaped by stigma, norms of masculinity, and limited opportunities for emotional disclosure, constraining help-seeking in offline settings. This study investigates how men use anonymous online peer-support spaces to discuss MI by analyzing discussions from the r/maleinfertility subreddit on Reddit. Using natural language processing techniques, we examined 10,769 posts and 80,381 comments published between 2013 and 2025. Analyses assessed sentiment and emotional expression, topic structure, hyperlink networks, and discussions related to diagnostic testing, treatment decision-making, and donor sperm use. Topic modeling revealed a functional differentiation between posts and comments. Original posts primarily focused on clinical sense-making, including interpretation of semen analyses, hormonal testing, and assisted reproduction options. In contrast, comments emphasized emotional validation, experiential knowledge-sharing, and normalization of alternative family-building pathways. Emotional expression varied by discussion topic, with heightened fear and sadness in conversations involving genetic testing, surgical sperm retrieval, and donor sperm. Hyperlink analysis indicated frequent engagement with peer-reviewed medical information, reflecting active evidence-seeking alongside peer exchange. Taken together, findings suggest that anonymous online communities function as critical infrastructures of support for men experiencing infertility, enabling forms of disclosure and vulnerability often constrained in offline contexts. These spaces facilitate interpretation of medical information, collective coping, and decision-making regarding treatment and donor options. The study highlights the role of digital anonymity in mitigating stigma and expanding communicative possibilities for men navigating infertility alongside clinical care.

## 1. Introduction

### 1.1 Clinical Complexity and Unmet Psychosocial Needs

Male infertility (MI) is a complex, multifactorial condition contributing to nearly half of all infertility cases worldwide and affecting 20–30% of reproductive-age couples (Agarwal et al., 2015; Huang et al., 2023; Li et al., 2025). Approximately 7% of men experience infertility, and chromosomal abnormalities account for roughly 4% of cases among the ∼2,000 genes involved in spermatogenesis (Houston *et al*., 2022; Mitrakas *et al*., 2025). While genetic defects explain approximately one-quarter of azoospermia cases, up to ∼40% of MI remains idiopathic, underscoring persistent gaps in etiological understanding (Krausz and Riera-Escamilla, 2018; Kuroda *et al*., 2020).

### 1.2 Diagnostic Testing, Treatment Decisions, and Donor Platform

Despite advances in molecular understanding, clinical evaluation of MI remains largely centered on medical history, physical examination, and WHO-standard semen analysis, with selective hormonal and genetic testing (Furini *et al*., 2025). Management strategies focus on lifestyle modification and correction of reversible causes, with Assisted Reproductive Technologies (ART) like Intrauterine Insemination (IUI), In vitro Fertilization (IVF), Intracytoplasmic Sperm Injection (ICSI) employed when conservative measures are unsuccessful (Schlegel *et al*., 2021a). However, the psychosocial dimensions of MI, including anxiety, decisional uncertainty, and identity concerns, remain comparatively underexplored.

Diagnostic investigations such as semen analysis, hormonal profiling, genetic testing, and, in selected cases, surgical sperm retrieval play a central role in guiding treatment decisions (Schlegel *et al*., 2021b, 2021a). Interpretation of these results is often complex, particularly in idiopathic or borderline cases, and can provoke substantial emotional distress that extends beyond the clinical encounter (Barratt *et al*., 2017). Treatment strategies are heterogeneous, with pharmacological interventions such as selective estrogen receptor modulators, gonadotropins, aromatase inhibitors, and antioxidant supplements frequently discussed in clinical practice and online forums, despite variable evidence and off-label use (Pallotti *et al*., 2022; Pozzi *et al*., 2024). Such uncertainties may shape men’s expectations, emotional responses, and perceptions of success or failure (Biggs *et al*., 2024; Sahoo *et al*., 2025).

For men with severe or irreversible spermatogenic failure, donor sperm offers a critical alternative, yet the decisions involve complex emotional, ethical, and identity-related considerations (Wang *et al*., 2025). Although donor conception has been widely examined from the perspectives of women undergoing treatment and intended parents more broadly, men’s own experiences and viewpoints remain poorly understood.

### 1.3 Psychosocial Burden, Gender Norms, and Digital Peer Support

Male infertility-related psychological stress has been linked to impaired semen parameters, suggesting bidirectional interactions between mental health and male reproductive function (Cao *et al*., 2025; Simbar *et al*., 2025). Although social media research has documented the psychosocial burden of infertility, existing studies disproportionately reflect female experiences, while male infertility discourse remains comparatively under-characterized despite evidence of active online engagement (Brochu et al., 2019). Sociocultural norms linking fertility to masculinity often discourage emotional disclosure and help-seeking, contributing to delayed care and unmet psychosocial needs (Dooley *et al*., 2011; Arya and Dibb, 2016; Patel *et al*., 2019). Consequently, men are increasingly turning to online communities for information and support, particularly since the COVID-19 pandemic has accelerated reliance on digital health resources (O’Connell *et al*., 2021; Sax and Lawson, 2022; Lin and Shorey, 2023). Beyond emotional support, online communities serve as a crucial source of medical information, although the quality and accuracy of the content vary widely. Analyses of male reproductive health discussions on Reddit reveal frequent discrepancies between peer-shared advice and clinical guidelines, highlighting gaps in understanding of diagnostic results and treatment options (Sellke *et al*., 2023).

### 1.4 Natural Language Processing and Study Rationale

Natural language processing (NLP) enables large-scale, longitudinal analysis of patient-generated text, capturing emotional tone, information needs, and behavioral patterns beyond the reach of surveys or clinic-based interviews (Hirschberg and Manning, 2015; Köckritz *et al*., 2025). Existing analyses have focused on specific sub-conditions such as hypogonadism, testosterone replacement therapy (TRT), varicocele, or small online cohorts, revealing recurrent themes of diagnostic confusion, unmet communication needs, and emotional distress (Osadchiy et al., 2020; Osadchiy, Mills and Eleswarapu, 2020; Sollender *et al*., 2024). To date, no study has integrated sentiment, emotion, topic modeling, and hyperlink analysis across a decade of male infertility discourse.

Addressing this gap, we applied an NLP framework to posts and comments from the r/maleinfertility subreddit spanning 2013–2025. This study was guided by four research questions: (RQ1) How are sentiment and emotional expression articulated and distributed within MI–related discussions? (RQ2) What key thematic patterns emerge across diverse male infertility–related discussions, and how do they influence user engagement and information sharing? (RQ3) What external information sources are shared in MI-related discussions, and how do these practices reflect users’ strategies for managing clinical uncertainty? (RQ4) How do emotional tone and discourse differ between posts and peer responses across clinical contexts? By integrating sentiment and emotion analysis, topic modeling, user engagement metrics, and hyperlink evaluation, this longitudinal study characterizes how men and their partners emotionally and cognitively navigate infertility outside clinical settings. In doing so, the study provides insights into unmet psychosocial and informational needs and highlights the potential of digital platforms to support more gender-equitable, patient-centered models of reproductive care.

## 2. Methodology

### 2.1 Study Design, Data Source, and Ethics

This study retrospectively analyzes longitudinal observational data derived from publicly accessible discussions on the r/maleinfertility subreddit on Reddit. The focus was exclusively on public content, with no attempts made to access private messages or to identify or contact users. Reddit usernames are unique for each user but do not reveal personal data; they were treated as anonymous identifiers. As the study relied exclusively on publicly available, de-identified data and involved no direct interaction with participants, institutional ethics review committee approval was not required. An overview of the methods used in the study can be found in Figure-1.

### 2.2 Data Collection and Eligibility

Posts and comments were obtained using the download utility from Project Arctic Shift (Heitmann, 2025) and exported in JSONL format. The data were then unpacked and converted into tabular files using Python (json and pandas). We included all posts and comments with non-empty text fields within the study window, excluding entries that were deleted or removed by moderators (e.g., labeled “[deleted]” or “[removed]”).

### 2.3 Text Preprocessing

In text-based analyses, the bodies of posts and comments were standardized by converting the text to lowercase and removing punctuation, email addresses, and hashtags. Common abbreviations were expanded, and stopwords (NLTK) were eliminated. To minimize noise in downstream models, spelling was corrected using the Python spellchecker library, and tokens were lemmatized with the NLTK WordNetLemmatizer. For hyperlink analyses, URL extraction was conducted on the original text before normalization.

### 2.4 User Activity and Engagement

User activity was tracked using the public, unique Reddit username as an anonymous identifier. We summarized this activity over time by counting posts, comments, and unique active users each month, defining an active user as someone who posted and/or commented in r/maleinfertility during that month.

### 2.5 Sentiment and Emotion Analysis

Sentiment was measured using the Valence Aware Dictionary and sEntiment Reasoner (VADER) from the NLTK package, which generated a sentiment score ranging from −1 to +1. These scores were classified as negative (≤ −0.05), neutral (−0.05 to +0.05), or positive (≥ +0.05) (Patel et al., 2026b). Emotions were identified using a pretrained model from HuggingFace by Hartmann (2022) that outputs Ekman’s six basic emotions (anger, disgust, fear, joy, sadness, surprise) along with neutral.

### 2.6 Theme Visualization and Topic Modeling

The wordcloud Python library (Mueller, 2020) was utilised to create word clouds, which visualize frequently occurring terms after the text was preprocessed and also after TF–IDF weighting was applied. To identify discussion themes, we applied BERTopic (Grootendorst, 2022). Top 10 topics were examined for both posts and comments. The Shifterator python package was used to generate a wordshift plot for the posts and comments of the subreddit [https://doi.org/10.1140/epjds/s13688-021-00260-3].

### 2.7 Hyperlink-based Analysis

Hyperlinks contained in posts and comments were extracted to characterize external information sources referenced by the community. Links were validated using Python’s requests library by confirming an HTTP 200 status code; only successful links were retained. We summarized link-sharing patterns by domain frequency to identify commonly cited sources.

### 2.8 Subreddit Cross-linking Analysis

Using Python’s re library, the regular expression pattern “/r/[a-zA-Z0-9_]+” was implemented to detect mentions of other subreddits. The extracted subreddit names were then verified for accuracy to remove any incorrect matches.

### 2.9 Keyword Analysis

A comprehensive dictionary of pharmacological agents, laboratory tests, and donor-related terms related to male infertility, along with their common synonyms and abbreviations, was compiled from the literature (Supplementary Tables 1, 2, and 3). Posts and comments were examined for these terms using case-insensitive, word-boundary matching. The occurrences were recorded, and the associated sentiment and emotion distributions were summarized for the matched texts. To evaluate whether the distribution of sentiments and emotions differed significantly across pharmacological agents, laboratory tests, or donor discussions, chi-square (χ²) tests of independence were performed using the corresponding contingency tables (Pearson, 1900). When expected cell counts were low, categories were combined as appropriate to satisfy test assumptions. Statistical significance was assessed using a two-sided α level of 0.05.

## 3. Results

### 3.1 Data Collection and Preprocessing

A total of 13,681 posts and 97,193 comments were collected from the r/maleinfertility subreddit. After pre-processing, the dataset contained 10,769 posts and 80,381 comments **(Table 1).** The number of posts and comments was tracked across the dataset. The subreddit had nominal activity until 2020, after which the activity increased over time, with a peak in October 2024 (Figure 2-A).

**Figure 1:**
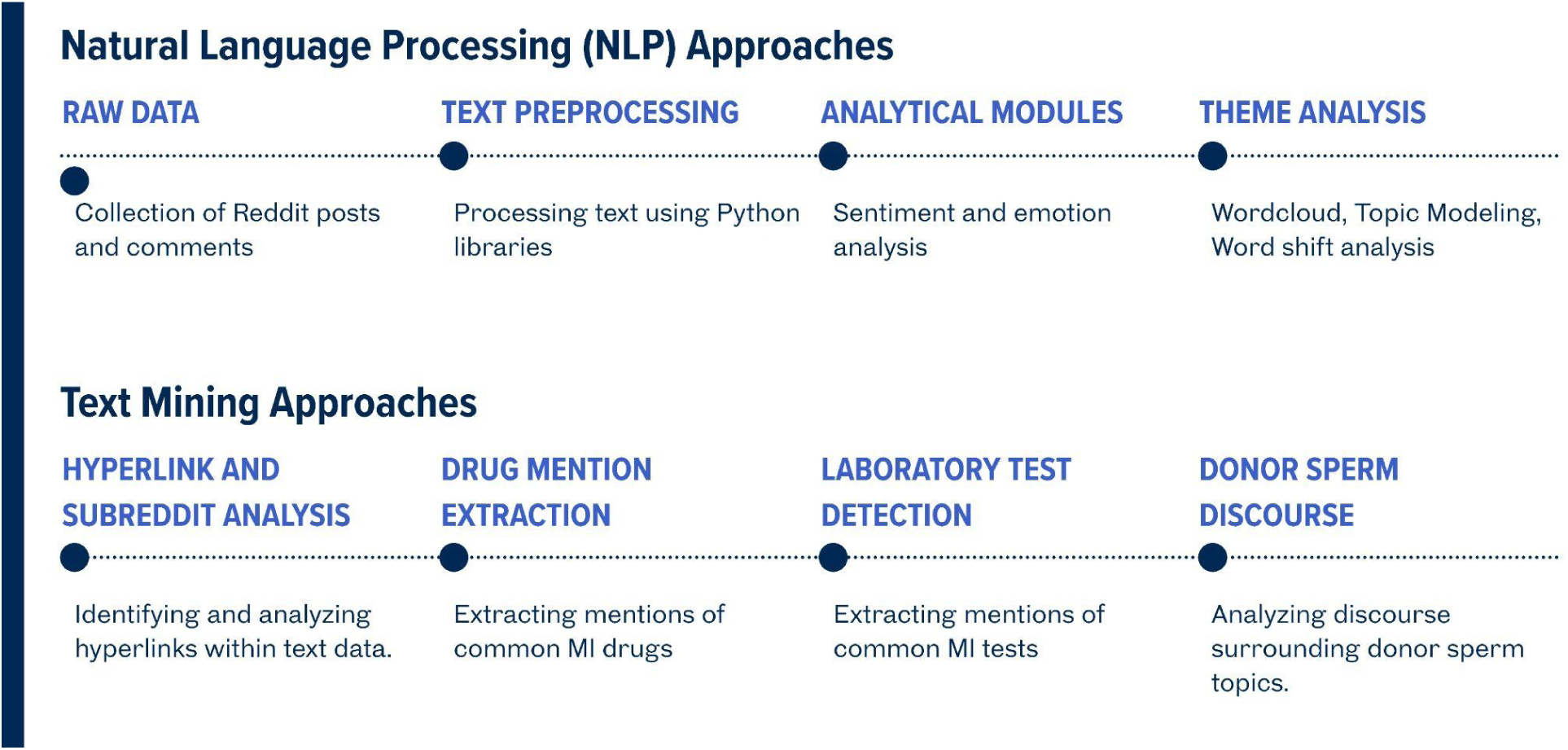
Overview of the methods and tools used in the study

**Figure 2.**
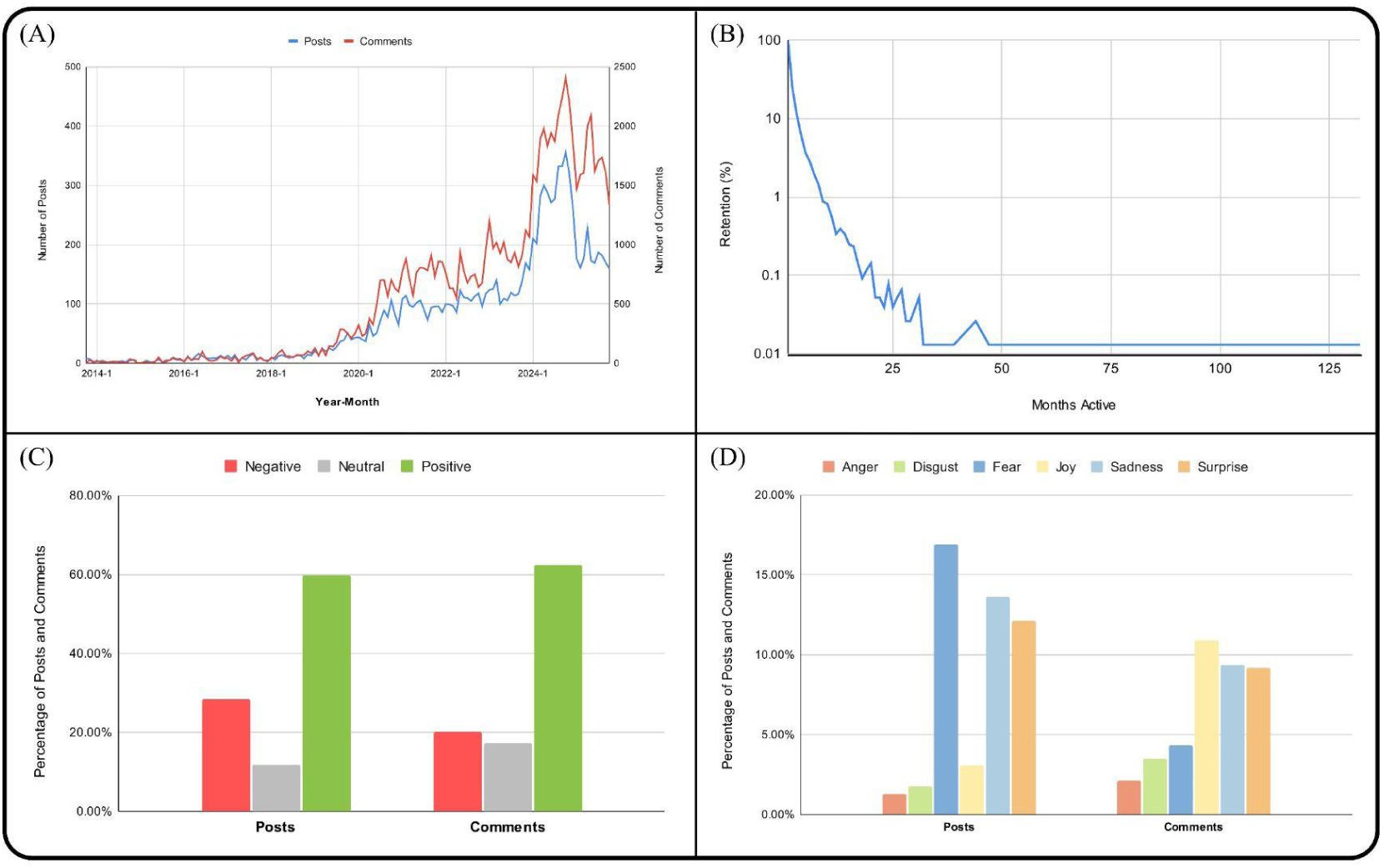
Overview of user engagement in the r/maleinfertility subreddit. (A) Distribution of sentiment (positive, neutral, and negative) expressed in posts and comments. (B) Distribution of emotions categories (anger, disgust, fear, joy, sadness, surprise) in posts and comments in the r/maleinfertility subreddit. (C) Temporal distribution showing the number of posts and comments per month. (D) User retention rate, illustrating continued participation over time in the r/maleinfertility subreddit.

**Table 1.** Dataset statistics of the collected r/maleinfertility subreddit data before and after pre-processing.

| Data | Collected | Deleted/Removed | Dataset |
| --- | --- | --- | --- |
| Posts | 13,681 | 2912 | 10,769 |
| **Comments** | 97,193 | 16,812 | 80,381 |
| **Total** | 110,874 | 19,724 | 91,150 |

**Table 2:** Examples of posts annotated as positive, negative, or neutral sentiments.

| Sentiment | Total posts | Examples |
| --- | --- | --- |
| Positive | 56633 (62.13%) | For anyone considering a varicocelelectomy to help with low sperm count, I would offer this encouraging news. My husband had the surgery about four months ago. 15 million/ml is considered normal, so from less than a million to almost normal is fantastic.<br>They found sperm! It's locked up, stored and on ice. The relief! Weird how I cannot recall ANY of the surgery. Thanks for all of your well wishes. :-D<br>This is really exciting! Obviously I wish you had sperm, but my fingers are crossed for you that you can make it happen. Feel free to message me if you have any questions, concerns, or wacky ideas. |
| Neutral | 15182<br>(16.66%) | Does the amount of days abstinent affect your count? Im guessing if Im 7 days vs 3 days my concentration would be higher in 7days?<br>The whole thing is a lot to think about.<br>We have a kit that we can send in the mail for people who cannot come into our clinic. All we need is a sperm sample (or just past test results) and a small blood sample or cheek swab. |
| Negative | 19335<br>(21.21%) | Hi Reddit... I just got my test results and the Dr said I have no sperm in my ejaculate. He had me give him a blood sample and another semen sample. I was wondering if anyone knows what I'm looking at. I have to call him at noon today, I'm just upset and disappointed.<br>Thanks. I am doing that currently. I am not currently on clomid though- so i can't blame outbursts on anything other than my own pain and sadness.<br>Now I'm worried about a tumor. You've listed everything we've eliminated, but that. I feelike Henny Penny: the sky is falling. |

### 3.2 User Activity

The temporal trends in posts and comments demonstrate a distinct correlation between the two (Figure 2-A). User activity remained relatively low until 2020, experienced an increase during the COVID-19 period, and then surged again in early 2024. Of the 12,048 users, 7,705 were one-time users, active for only a month. This was considered 100% retention, and a graph was plotted illustrating months active versus retention (Figure 2-B). The retention curve exhibited a steep initial decline, followed by a slight leveling off and then followed by a flat tail.

### 3.3 (RQ1) How are sentiment and emotional expression articulated and distributed within MI–related discussions?

#### 3.3.1 Sentiment Analysis

Overall sentiment analysis indicates that both posts and comments are predominantly positive (Figure 2-C). Posts exhibit a majority of positive sentiment (59.71%), followed by negative sentiment (28.53%), which exceeds the proportion of neutral sentiment (11.77%). Similarly, comments are largely positive (62.46%), while negative (20.23%) and neutral (17.31%) sentiments occur at similar and low levels. Table-2 shows the number, percentage, and examples of posts and comments categorized by sentiment type.

#### 3.3.2 Emotion Analysis

Emotion analysis reveals distinct patterns between posts and comments (Figure 2-D). In posts, fear is the most dominant emotion (16.89%), followed by sadness (13.63%) and surprise (12.08%). In contrast, emotions such as joy (3.06%), anger (1.29%), and disgust (1.75%) are minimally represented. For comments, joy emerged as the most prominent emotion (10.92%), followed by sadness (9.35%) and surprise (9.20%). Fear (4.34%), anger (2.08%), and disgust (3.46%) are present at relatively low levels.

#### 3.3.3 RQ1 Conclusion

Discussions related to MI are positive in sentiment, with most posts and comments expressing favorable views. However, posts contain a relatively higher proportion of negative sentiment, indicating that users often share concerns and emotional difficulties. In contrast, comments tend to be more supportive and affirming. Emotion analysis shows that posts are mainly driven by fear and sadness, reflecting anxiety and uncertainty. Comments, however, are more strongly associated with joy and moderate levels of sadness and surprise, suggesting empathetic and encouraging responses.

### 3.4 (RQ2) What key thematic patterns emerge across diverse male infertility–related discussions, and how do they influence user engagement and information sharing?

#### 3.4.1 Theme and Topic Modeling

The wordcloud revealed that some words, like doctor, test, results, and sperm, were prevalent in the posts (Figure 3-A). While husband, doctor, test, and results were common in the comments (Figure 3-B). The TF–IDF-weighted word clouds for posts and comments showed distinct patterns in discussions of male infertility. Specifically, in posts, prominent terms include “test”, “sperm”, “motility”, “count”, and “morphology”. Moreover, medical terms such as “urologist”, “varicocele”, and “azoospermia” are also present. In addition, relational and temporal terms including “wife”, “partner”, “month”, and “year” appear frequently (Figure 3-C). In comments, frequently occurring terms include “thanks”, “luck”, “hope”, “help”, and “best”. Terms related to clinical indicators and treatments, such as “testosterone”, “surgery”, “supplement”, and “count”, are also observed (Figure 3-D).

**Figure 3.**
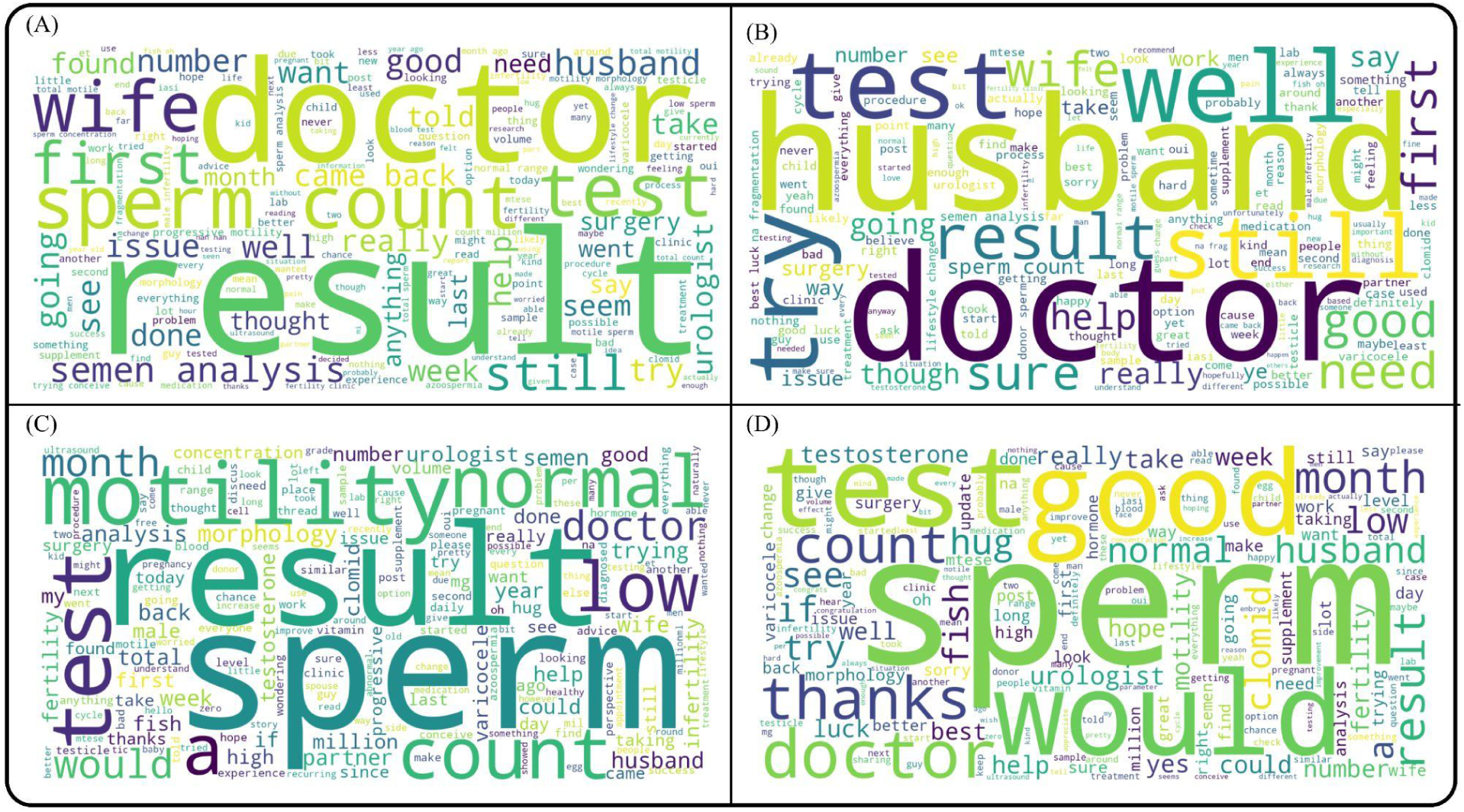
Word clouds generated using the (A) Posts and (B) Comments (C) TF-IDF Posts and (D) TFIDF comments

Topic modeling revealed that the most dominant topic in the posts is IVF and semen analysis, with users frequently discussing their diagnostic test results and seeking advice or support from the subreddit. The second most prominent topic focuses on treatment options for male infertility, followed by discussions related to consultations with urologists. In contrast, the most discussed topic in the comments is emotional support and gratitude, where users encourage post authors and offer reassurance to help them remain hopeful and look for alternatives. The second major topic concerns donor sperm and adoption, which are commonly discussed alternatives for individuals and couples facing MI. The third prominent topic in the comments relates to treatment options, including discussions of the various medications currently available. The top 10 topics for posts and comments are presented in **Table 3**.

**Table 3.** Top 10 topics extracted from posts and comments in the r/male infertility subreddit.

| Posts |  |  |
| --- | --- | --- |
| Topic Number | Count | Description |
| 1 | 3569 (38.7%) | IVF and semen analysis |
| 2 | 858 (9.31%) | Hormonal male infertility treatment |
| 3 | 685 (7.42%) | Urologist and male infertility |
| 4 | 400 (4.34%) | Semen analysis metrics |
| 5 | 314 (3.41%) | Low sperm and testosterone |
| 6 | 303 (3.29%) | Clomiphene for male infertility |
| 7 | 270 (2.93%) | Improving sperm quality |
| 8 | 252 (2.73%) | Testicular health issues |
| 9 | 242 (2.62%) | Donor sperm conception |
| 10 | 228 (2.47%) | hCG and Clomid therapy |
| Comments |  |  |
| Topic Number | Count | Description |
| 1 | 36852 (45.85%) | Emotional support and gratitude |
| 2 | 3396 (4.22%) | Donor sperm and adoption |
| 3 | 3177 (3.95%) | Male infertility and treatment |
| 4 | 1628 (2.03%) | IVF and IUI outcomes |
| 5 | 1198 (1.49%) | Success stories and congratulations |
| 6 | 1096 (1.36%) | Recovery and well-wishes |
| 7 | 1070 (1.33%) | Semen analysis parameters |
| 8 | 854 (1.06%) | Urologist consultations |
| 9 | 854 (1.06%) | IVF decisions and experiences |
| 10 | 743 (0.92%) | Testicular health concerns |

**Table 4.** Top 10 hyperlinks extracted from posts and comments in the r/maleinfertility subreddit.

| Domain | Posts | Comments | Total |
| --- | --- | --- | --- |
| https://www.reddit.com | 193 | 480 | 673 |
| https://preview.redd.it | 196 | 161 | 357 |
| https://pmc.ncbi.nlm.nih.gov | 57 | 224 | 281 |
| https://pubmed.ncbi.nlm.nih.gov | 34 | 185 | 219 |
| https://np.reddit.com | 0 | 208 | 208 |
| https://imgur.com | 82 | 64 | 146 |
| https://www.amazon.com | 4 | 96 | 100 |
| https://www.youtube.com | 10 | 70 | 80 |
| https://i.imgur.com | 34 | 29 | 63 |
| https://www.fertstert.org | 18 | 39 | 57 |

The word-shift analysis comparing r/maleinfertility posts and comments reveals clear differences in language use between the two. Posts are dominated by medically oriented and self-focused terms, such as “result,” “motility,” “normal,” “sperm,” “concentration,” “count,” “analysis,” and “morphology.” In contrast, comments are characterized by supportive, emotional, and encouraging language. Words such as “good,” “best,” “luck,” “yes,” “sorry,” “hug,” “hope,” “well,” “definitely,” and “would” show positive shift scores, indicating higher usage in comments (Supplementary Figure-1).

#### 3.4.2 RQ2 Conclusion

Word-cloud, TF–IDF, and word-shift analyses further confirm this distinction, with posts dominated by technical and self-focused terms and comments characterized by supportive and empathetic language. Topic modeling results show clear functional differences between posts and comments in MI-related discussions. Posts are primarily focused on medical evaluations, diagnostic results, and treatment options, reflecting users’ information-seeking behavior and personal health concerns. In contrast, comments emphasize emotional support, encouragement, and alternative pathways such as donor sperm and adoption.

### 3.5 (RQ3) What external information sources are shared in MI-related discussions, and how do these practices reflect users’ strategies for managing clinical uncertainty?

#### 3.5.1 Hyperlink-based Analysis

The most common hyperlinks were crosslinks to Reddit (www.reddit.com) and the domain where any uploaded photos are stored (<u>preview.redd.it</u>). But the hyperlink analysis revealed that the subreddit shared more peer-reviewed research articles from PubMed (<u>pmc.ncbi.nlm.nih.gov</u>, <u>pubmed.ncbi.nlm.nih.gov</u>, www.fertstert.org/) when compared to unreliable news articles. The www.np.reddit.com is interesting, the “NP” stands for “No Participation”, its intended use is to prevent brigading (people from one sub coming to another they’re not a part of and interfering). Imgur (<u>imgur.com</u>, <u>i.imgur.com</u>) is also an image-sharing platform, and people can upload any image without censoring and share the link on Reddit. The Amazon (www.amazon.com) links are used to share useful products, and YouTube (www.youtube.com/) is used to share videos. The hyperlinks along with their occurrence in posts and comments can be found in Table 5 and Supplementary Table 4.

**Table 5.** Top 10 referenced subreddits extracted from posts and comments in the r/maleinfertility subreddit.

| Subreddit | Posts | Comments | Total |
| --- | --- | --- | --- |
| /r/infertility | 18 | 46 | 64 |
| /r/dnafragmentation | 0 | 20 | 20 |
| /r/donorconceived | 0 | 13 | 13 |
| /r/guyvf | 0 | 13 | 13 |
| /r/maleinfertility | 6 | 6 | 12 |
| /r/askadcp | 0 | 10 | 10 |
| /r/cryo_kids | 0 | 9 | 9 |
| /r/varicocele | 2 | 5 | 7 |
| /r/tryingforababy | 2 | 5 | 7 |
| /r/trollingforababy | 0 | 5 | 5 |

#### 3.5.2 Subreddit Analysis

The users of the r/male infertility subreddit cross-reference the r/infertility subreddit the most, followed by r/dnafragmentation, /r/donorconceived, and /r/guyvf. The list of subreddits, along with the count in posts and comments, can be found in Table 5 and Supplementary Table 5.

#### 3.5.3 RQ3 Conclusion

The hyperlink analysis shows that users in the r/maleinfertility subreddit primarily rely on credible and scientifically validated sources, particularly peer-reviewed articles from PubMed and established medical journals. In addition, frequent crosslinks to Reddit and related subreddits indicate the importance of peer communities for sharing experiences and contextualizing medical knowledge. Users share images, videos, and shopping links to supplement their opinions and advice with visual evidence, practical examples, and recommended resources.

### 3.6 (RQ4) How do emotional tone and discourse differ between posts and peer responses across clinical contexts?

#### 3.6.1 Drug Mention Analysis

A total of 11 classes of drugs were identified from the literature as potential treatments for MI. Of these, the drug classes “Clomiphene citrate,” “Letrozole, Anastrozole,” “CoQ10, L-carnitine, vitamins C/E, zinc, selenium, NAC,” and “Exogenous testosterone (TRT)” exceeded the predefined activity thresholds (>100 mentions in posts and >1000 mentions in comments). Sentiment analysis of both posts and comments revealed that positive sentiment was predominant, accounting for more than 60% of the content (Figure 4-A,B). Negative sentiment was comparatively lower, while neutral sentiment constituted only a small proportion. Consistent with these observations, Pearson’s chi-square tests indicated a significant association between drug class and sentiment distribution in both posts (χ²(6) = 152.94, p = 1.85 × 10⁻³⁰) and comments (χ²(6) = 320.89, p = 2.72 × 10⁻⁶⁶). Emotion analysis indicated that neutral emotion was the most dominant overall (Supplementary Figure-2). Excluding neutral emotion, fear, sadness, and surprise were the most prevalent emotions in posts (Figure 4-C). A significant association between drug class and emotional tone was observed for both posts (χ²(18) = 52.40, p = 3.25 × 10⁻⁵) and comments (χ²(18) = 401.16, p = 5.25 × 10⁻⁷⁴). In contrast, sadness was the dominant emotion in comments across most drug classes, except for discussions involving CoQ10, L-carnitine, vitamins C/E, zinc, selenium, and N-acetylcysteine (NAC), where levels of sadness and anger were comparatively lower (Figure 4-D).

**Figure 4.**
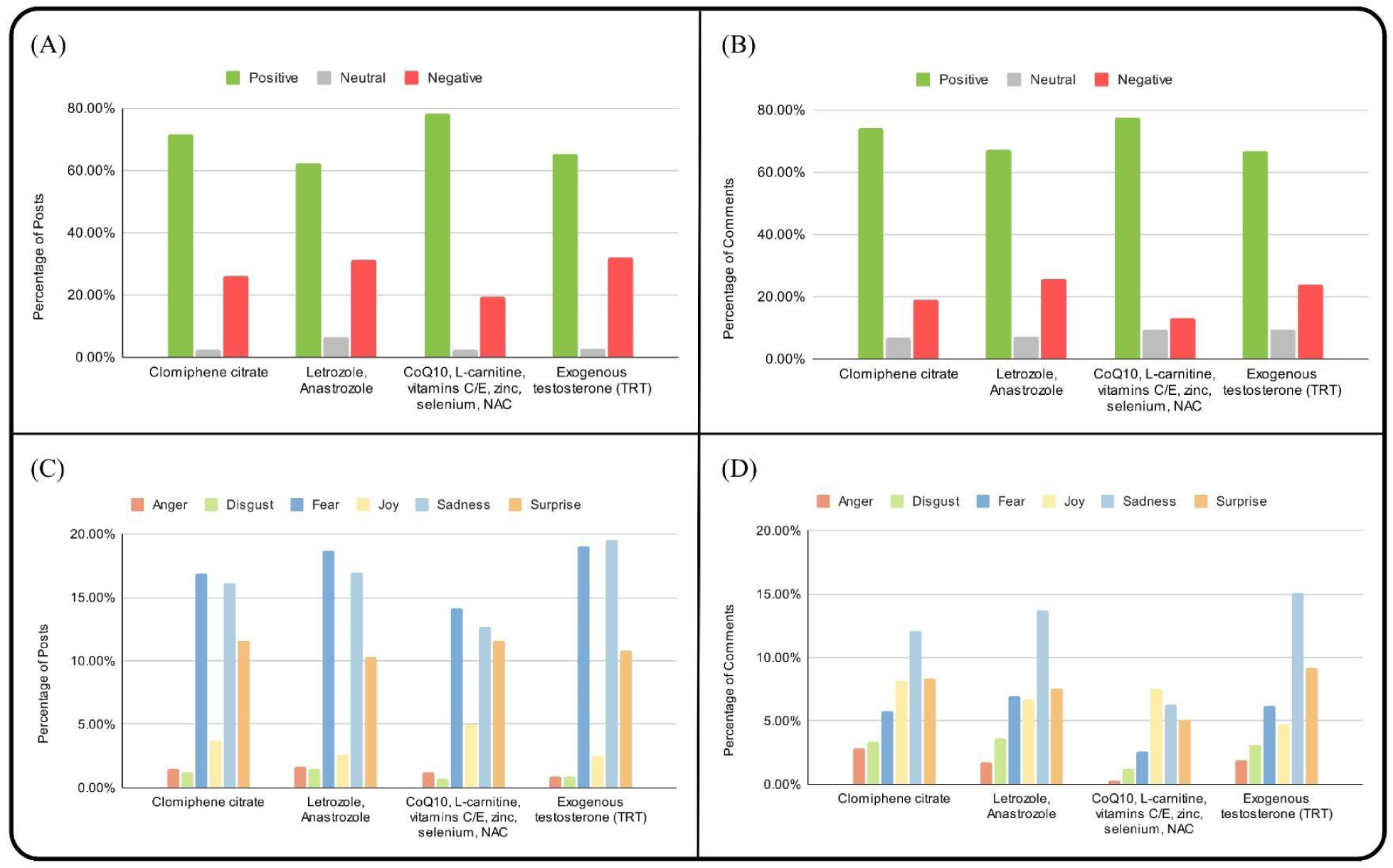
Analysis of posts and comments discussing drugs in the r/maleinfertility subreddit. (A) Distribution of sentiment (positive, neutral, and negative) in posts. (B) Distribution of emotion categories (anger, disgust, fear, joy, sadness, and surprise) expressed in posts. (C) Distribution of sentiment (positive, neutral, and negative) in comments. (D) Distribution of emotion categories (anger, disgust, fear, joy, sadness, and surprise) expressed in comments.

#### 3.6.2 Laboratory Test Analysis

A total of 10 types of tests were identified from the literature as potential treatments for MI. Of these, six tests exceeded the predefined activity thresholds (>100 mentions in posts and >1000 mentions in comments). Sentiment analysis of both posts and comments revealed that positive sentiment was predominant, accounting for 58%-70% of the content (Figure 5-A,B). Negative sentiment was comparatively lower, ranging from 22% to 40%, while neutral sentiment constituted only a small proportion. Consistent with these trends, Pearson’s chi-square tests demonstrated a significant association between test type and sentiment distribution in both posts (χ²(10) = 47.44, p = 7.86 × 10⁻⁷) and comments (χ²(10) = 87.68, p = 1.54 × 10⁻¹⁴). Emotion analysis indicated that neutral emotion was the most dominant overall (Supplementary Figure-3). Excluding neutral emotion, fear, sadness, and surprise were the most prevalent emotions in posts (Figure 5-C). A significant association between laboratory test type and emotional tone was also observed for both posts (χ²(30) = 61.24, p = 6.49 × 10⁻⁴) and comments (χ²(30) = 578.97, p = 6.65 × 10⁻¹⁰³). In comments, fear and sadness were dominant emotions, with fear being very high in “genetic Testing” and “Testicular Biopsy / Micro-TESE” (Figure 5-D).

**Figure 5.**
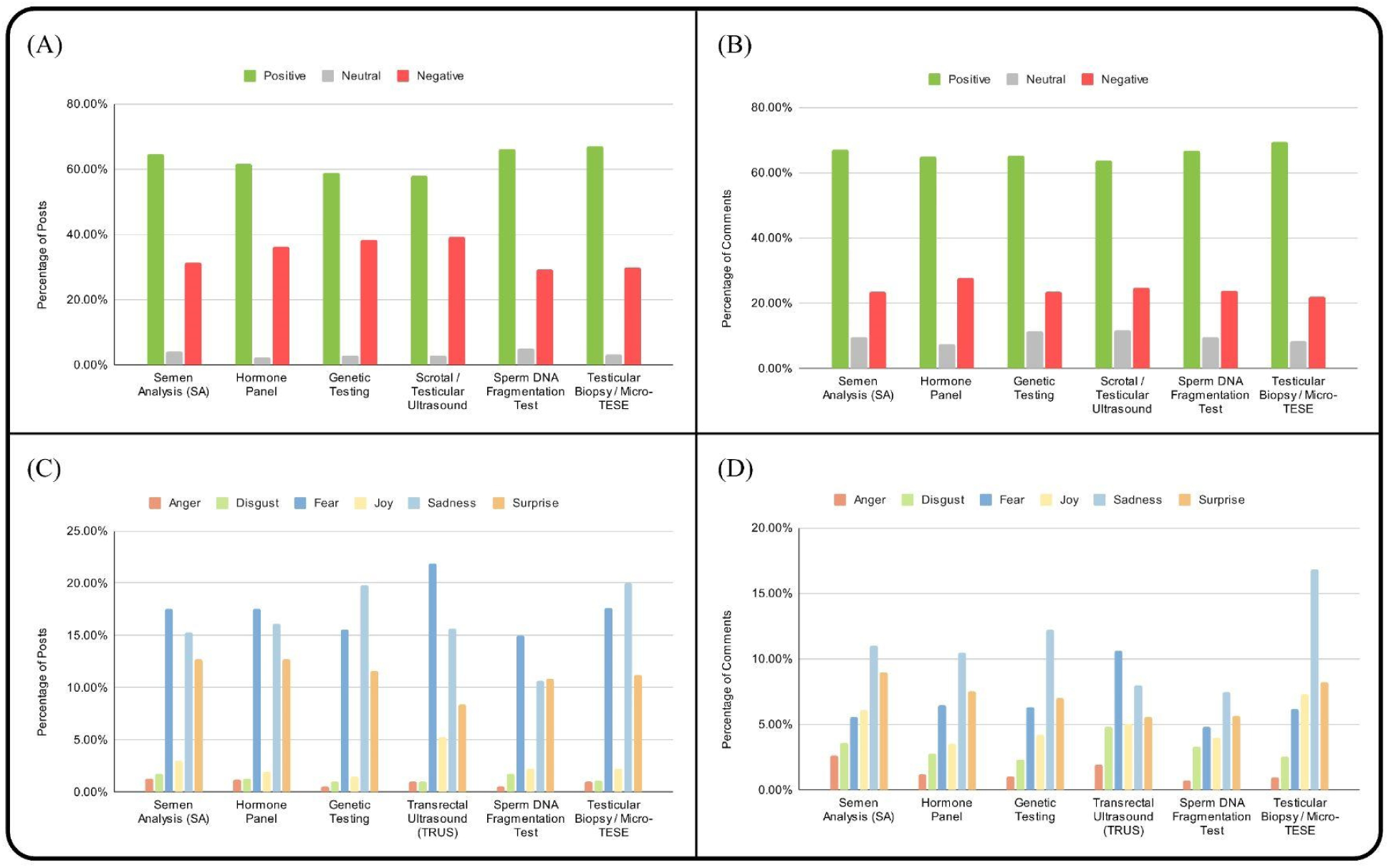
Analysis of posts and comments discussing prescribed tests in the r/maleinfertility subreddit. (A) Distribution of sentiment (positive, neutral, and negative) in posts. (B) Distribution of emotion categories (anger, disgust, fear, joy, sadness, and surprise) expressed in posts. (C) Distribution of sentiment (positive, neutral, and negative) in comments. (D) Distribution of emotion categories (anger, disgust, fear, joy, sadness, and surprise) expressed in comments.

#### 3.6.3 Donor Discussion Analysis

Three sets of keywords corresponding to Donor Sperm (General), Sperm Banks and Procurement, and Clinical Use of Donor Sperm were identified from the literature related to MI. Sentiment analysis of both posts and comments revealed that positive sentiment was predominant, with comments exhibiting slightly higher positivity than posts (Figure 6-A,B). Consequently, negative sentiment was comparatively lower in comments than in posts, while neutral sentiment accounted for only a small proportion of the content. However, Pearson’s chi-square tests indicated no significant association between donor-related topic and sentiment distribution in either posts (χ²(4) = 1.54, p = 0.82) or comments (χ²(4) = 8.23, p = 0.084). Emotion analysis indicated that neutral emotion was the most dominant overall (Supplementary Figure-4). Excluding neutral emotion, fear, sadness, and surprise were the most prevalent emotions expressed in posts (Figure 6-C), although no significant association between donor topic and emotional tone was observed for posts (χ²(12) = 14.64, p = 0.26). In contrast, emotional tone differed significantly across donor-related categories in comments (χ²(12) = 62.01, p = 9.70 × 10⁻⁹). In the comments, fear was present across all three categories and was particularly dominant in discussions related to ‘Donor Sperm (General)’. Surprise was notably associated with discussions on ‘Sperm Banks and Procurement’, while joy was consistently present across all three categories, reflecting supportive and reassuring community interactions (Figure 6-D).

**Figure 6.**
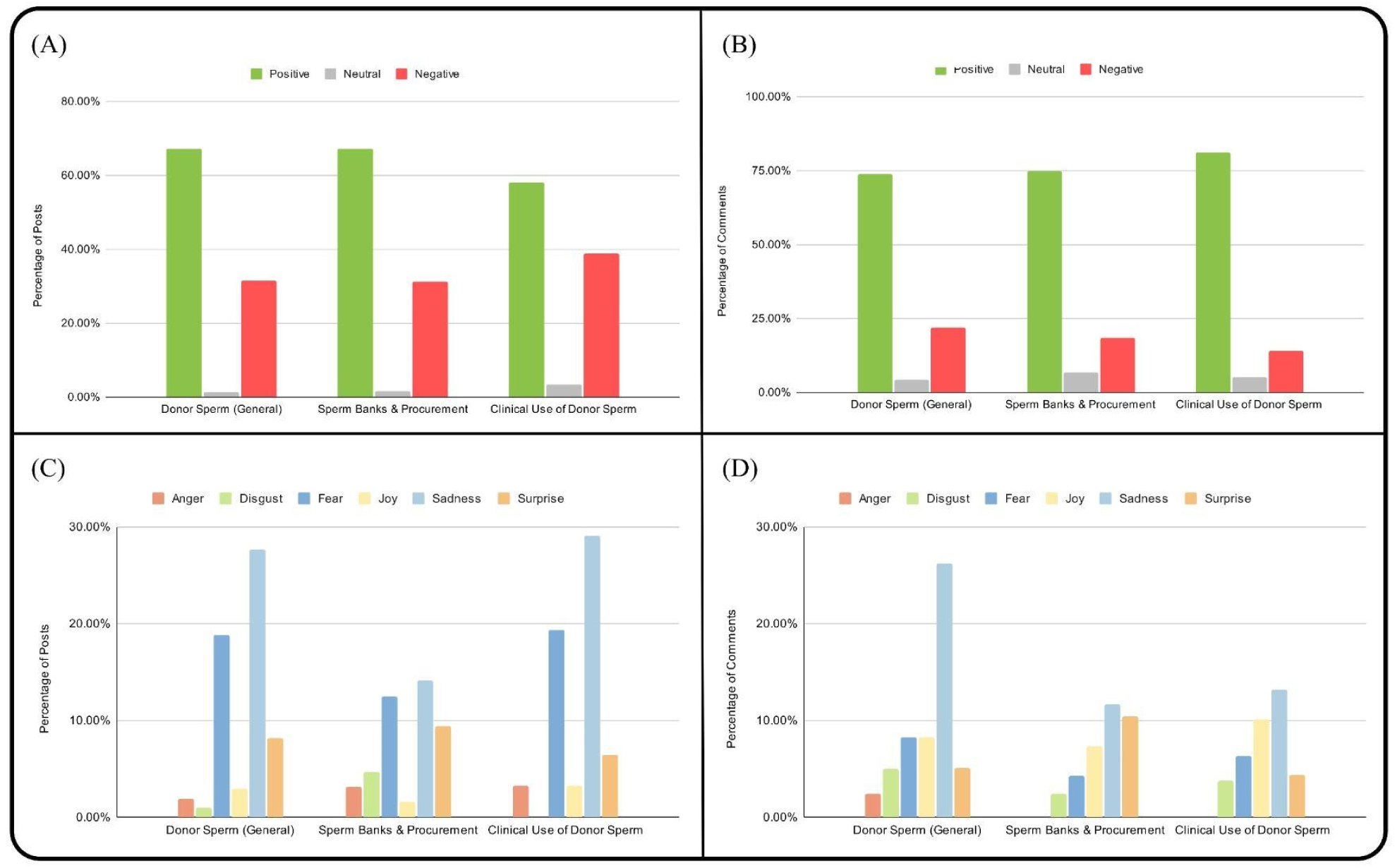
Analysis of posts and comments discussing sperm donors in the r/male infertility subreddit. (A) Distribution of sentiment (positive, neutral, and negative) in posts. (B) Distribution of emotion categories (anger, disgust, fear, joy, sadness, and surprise) expressed in posts. (C) Distribution of sentiment (positive, neutral, and negative) in comments. (D) Distribution of emotion categories (anger, disgust, fear, joy, sadness, and surprise) expressed in comments.

#### 3.6.4 RQ4 Conclusion

The analysis demonstrates that emotional tone and discourse differ systematically between posts and peer responses across clinical contexts. Across drug, laboratory test, and donor-related discussions, positive sentiment consistently dominates both posts and comments, indicating an overall supportive communication environment. However, posts are more frequently characterized by fear, sadness, and surprise, reflecting users’ uncertainty, anxiety, and emotional vulnerability when discussing treatments, diagnostic procedures, and reproductive options.In contrast, comments tend to exhibit higher levels of sadness and, in some contexts, joy. Significant associations between clinical categories and emotional tone in most analyses is identified, particularly in discussions of pharmacological treatments and invasive diagnostic tests. Notably, donor-related discussions show more stable sentiment patterns in posts but greater emotional variation in comments.

## 4. Discussion

This study aimed to provide a longitudinal characterization of how men discuss, interpret, and emotionally process MI within a large, male-focused online community. By integrating sentiment and emotion analysis with topic modeling, hyperlink extraction, and focused examinations of medical treatment, diagnostic testing, and donor-related discourse, we identify a consistent pattern of diagnostic anxiety in help-seeking posts and affective buffering through comments. Our findings extend prior qualitative research on infertility-related distress and help-seeking behaviors (Fisher and Hammarberg, 2012; Sahoo et al., 2025) by demonstrating, how men navigate uncertainty, masculinity, and decision-making outside formal clinical settings.

Addressing RQ1, sentiment and emotional expression in MI-related discussions reveal a stable asymmetry over time, with posts dominated by fear, sadness, and surprise during diagnostic and decisional moments, and comments exhibiting more positive emotional tones that reflect reassurance and peer support. In response to RQ2, thematic analyses show a clear functional division in which posts primarily focus on diagnostic testing, treatment decision-making, and donor sperm, while comments emphasize emotional labor, experiential knowledge, and normalization. With respect to RQ3, hyperlinking practices indicate active evidence-seeking strategies, characterized by frequent sharing of peer-reviewed medical sources and structured subreddit cross-linking, reflecting an effort to minimize misinformation. Finally, addressing RQ4, emotional tone and discourse differ markedly between posts and comments, particularly in donor-related contexts, where expressions of distress and identity threat in posts are met with affirming, hopeful, and acceptance-oriented responses in comments.

### 4.1 Growth, Engagement, and Episodic Use of Digital Peer Support

Our data demonstrates that community activity was modest from inception until 2020, followed by sustained growth that peaked in October 2024, a pattern consistent with broader post–pandemic shifts toward online peer support and tele–reliant information–seeking in infertility care. Building on prior work showing increased online engagement among infertility communities after COVID–19 (Morgan *et al*., 2020), our temporal trend supports the view that Reddit functions as an adaptive extension of the clinic for MI (Patel *et al*., 2019).

Despite increasing activity, retention analyses reveal a pronounced high–churn/low–core structure, with most users participating only for a single month, producing a steep early drop–off with a shallow long–tail of repeat contributor. This configuration is typical of communities that serve users at discrete inflection points (e.g., new diagnosis, abnormal semen analysis, or treatment failure), while a small subset of repeat contributors offer experiential guidance and emotional continuity (Carron-Arthur *et al*., 2014).

### 4.2 Emotional Asymmetry Between Help-Seeking and Help-Giving

Although aggregate sentiment was predominantly positive across both posts and comments, emotion analysis revealed a consistent asymmetry. Posts were dominated by fear, sadness, and surprise, whereas comments showed higher levels of joy, reassurance, and gratitude. This divergence reflects a classic help-seeking/help-giving dynamic observed in other online health communities in infertility and the buffering role of social support (Fisher and Hammarberg, 2012).

Posts were marked by a heightened emotional complexity, with fear, sadness, and surprise being the predominant emotions expressed. This aligns with previous findings on the emotional burden and anticipatory stress linked to the diagnostic uncertainty associated with infertility (Boivin et al., 2023; Lin and Shorey, 2023). These emotional surges may stem from the demands of traditional diagnostic testing, the interpretation of semen analysis results, and the consideration of invasive procedures like testicular biopsy or micro-TESE during the MI journey (Hanna and Gough, 2016a; Hanna and Gough, 2016b; Hanna, Gough and Hudson, 2018). In contrast, the relative emotional positivity of comments underscores the buffering role of the community. This is supported by the word shift analysis which identified a distinct linguistic divergence between original posts and their subsequent comments. Posts were characterized by a significant shift toward diagnostic and biometric tokens, with high-contribution terms including “motility,” “concentration,” “morphology,” “count,” and “analysis.” This technical lexicon highlights a focus on clinical data and laboratory results. Conversely, comments shifted toward an affective and supportive lexicon, dominated by tokens such as “luck,” “sorry,” “hope,” “best,” and “definitely.” These findings align with evidence that men experiencing infertility often struggle with disclosure and receive disproportionate emotional support from the public (Fisher and Hammarberg, 2012). For comparison, a recent meta-ethnographic review based on face-to-face interviews identified emotional suppression as a dominant response among men with infertility (Abbasi et al., 2025). Our findings suggest that anonymous online interactions may enable greater emotional disclosure and provide psychological relief through positive peer response, partially offsetting gaps in formal care.

### 4.3 Thematic Polarization: Clinical Sense-Making Versus Emotional Labor

Topic modeling further clarifies this functional division. Posts center on diagnostic interpretation and decisions, whereas comments emphasize emotional support, donor/adoption considerations, and practical next steps. This division underscores a functional specialization within the community: users initiate discussions to seek interpretation of clinical data and guidance on next steps, while responders provide emotional labor, encouragement, and experiential insight (Holley *et al*., 2015).

Word-frequency patterns reinforce this distinction, with posts emphasizing clinical nouns (“doctor,” “test,” “results,” “sperm”) that signal active interpretation of biomedical data. Collectively, these results position the subreddit as a distributed interpretive space where biomedical information is socially processed rather than passively consumed (Osadchiy, Mills and Eleswarapu, 2020).

### 4.4 Information Pathways and Quality of Shared Resources

Hyperlink analysis revealed a notable preference for peer-reviewed and government sources, like PubMed, Fertility and Sterility journal, and ClinicalTrials.gov, suggesting relatively strong engagement with primary literature compared with purely commercial or news sources. This pattern contrasts with concerns that social media health discussions are primarily driven by misinformation, suggesting that this community exhibits relatively high scientific literacy (Treadgold *et al*., 2025; Patel *et al*., 2026a).

The frequent use of np.reddit.com links suggests community familiarity with Reddit’s “no participation” convention, often employed when referencing external discussions to reduce direct interaction and maintain separation in sensitive and personal conversations. Rather than discouraging online engagement, clinicians may benefit from acknowledging these platforms and proactively directing patients towards validated, evidence-based resources to help address uncertainty and clarify misconceptions.

### 4.5 Treatment Discourse: Optimism Modulated by Perceived Invasiveness

Across the drug classes, sentiment remained predominantly positive in both posts and comments, reflecting optimism toward both medical and lifestyle-based interventions. In contrast, emotions reveal nuance: fear, sadness, and surprise concentrate around hormonal therapies and more invasive trajectories. In contrast, nutraceutical discussions (e.g., CoQ10, L–carnitine, antioxidants) elicit comparatively less negative affect (Zafar *et al*., 2023). With Chi-square results confirming significant heterogeneity, this gradient suggests that perceived invasiveness, irreversibility, and cost shape emotional responses. These findings underscore the importance of anticipatory counseling that explicitly addresses emotional as well as physiological implications of the different treatment options (Sahoo *et al*., 2025).

### 4.6 Diagnostic Testing as a Psychological Inflection Point

Although sentiment surrounding diagnostic testing was generally positive, fear and sadness peaked in discussions around genetic testing and testicular biopsy/micro–TESE, commonly used for idiopathic MI diagnosis (Olesen *et al*., 2017). These procedures often represent decisive moments that redefine reproductive possibilities and confront men with potential limits to biological parenthood. The emotional intensity observed at these junctures highlights the need for targeted psychological support and clearer pre-test counseling to reduce uncertainty and decisional regret (Biggs *et al*., 2024). Consistent with these observations, chi-square analyses indicated that sentiment and emotional tone differed significantly across diagnostic test categories, particularly within comments.

### 4.7 Donor Sperm Discussions and Identity Transition

Donor sperm discourse emerged as a distinct thematic and emotional domain (Burr, 2010; Zadeh *et al*., 2016). Posts initiating donor-related discussions were characterized by elevated fear and sadness, reflecting grief over genetic loss, concerns about masculinity, and relational implications (Kalampalikis *et al*., 2018). Quantitatively, however, these emotional patterns in posts were broadly similar across donor-related categories, as reflected by the absence of a significant association between donor topic and emotional tone. This suggests that initial discussions of emotional responses to donor sperm are largely shared across donor-related contexts rather than being topic-specific.

In contrast, comments frequently expressed joy, reassurance, and normalization, emphasizing acceptance and successful outcomes. Consistent with this qualitative shift, chi-square analysis revealed a significant association between donor-related category and emotional tone in comments, indicating that emotional responses differed systematically by specific donor-related context. The findings position donor decision-making as a critical psychosocial inflection point where peer support may be particularly impactful and may play an essential role in facilitating emotional adjustment and reducing stigma (Hanna *et al*., 2018).

### 4.8 Clinical and Research Implications

Collectively, these findings suggest several actionable implications. First, clinicians should anticipate and address patients’ diagnostic sense-making needs through clear, plain-language explanations of semen analysis results, treatment pathways, and decision thresholds. Second, because high early churn suggests users arrive during acute stress points and leave once immediate needs are resolved, support should be timed to these moments through short, modular interventions like two-visit “diagnosis and decisions” bundles, brief digital handouts, and curated lists of trusted links aligned with testing, procedures, and treatment transitions. Third, given the community’s strong engagement with primary literature, clinician-curated digital resources and guidance on evaluating supplements and commercial claims may enhance informed decision-making rather than compete with online discourse.

Methodologically, this study demonstrates the value of integrating sentiment analysis, emotion classification, topic modeling, and hyperlink mapping to capture both informational and affective dimensions of patient discourse at scale, complementing traditional qualitative approaches.

### 4.9 Strengths and Limitations

Strengths of this study include its longitudinal scope and robust NLP framework and focus on a male-specific infertility forum where men’s experiences are often underrepresented. However, several limitations should be acknowledged. Reddit users are not geographically or demographically representative of all men experiencing infertility, as the platform is predominantly used in developed countries. Additionally, the anonymous nature of Reddit prevents verification of individual-level demographic characteristics, such as age and gender, as well as any declared clinical diagnoses or treatment outcomes. The analysis is restricted to English-language content from a single platform, while lexicon-based sentiment methods can oversimplify or miss irony and sarcasm. In addition, activity and retention metrics reflect visible participation rather than passive engagement, and hyperlink counts serve only as rough proxies for information quality rather than definitive measures. Future work can be expanded to encompass other social media platforms such as Facebook, X, or YouTube. Additionally, further research can incorporate other data modalities, including images, videos, and documents shared by users on these platforms.

## 5. Conclusions

In conclusion, this study provides the first longitudinal, computationally driven examination of MI discourse within an online peer-support community. The findings reveal a dynamic interplay between diagnostic anxiety, treatment optimism, and peer-mediated emotional support, highlighting how men actively process infertility-related uncertainty, decision-making, and emotional distress outside traditional clinical settings. Importantly, the results underscore a critical opportunity to normalize MI within public and clinical narratives through gender-equitable engagement that complements rather than detracts from the well-established burden of female infertility. By utilizing trusted digital platforms, improving clinician–patient communication, and encouraging earlier peer-supported help-seeking, insights from this study can inform more inclusive, couple-centered models of reproductive care and strengthen the role of digital communities in male reproductive health.

## Data availability statement

Publicly available datasets were analyzed in this study. This data can be found at: https://www.reddit.com/.

## Data Availability

Publicly available datasets were analyzed in this study. This data can be found at: https://www.reddit.com/ and downloaded using https://github.com/ArthurHeitmann/arctic_shift.

## References

Abbasi, F., Talebi, M., Jandaghian-Bidgoli, M., Shaterian, N. and Dehghankar, L. (2025). Surviving and thriving: male infertility through the lens of meta-ethnography. Discover Social Science and Health, 5(1). 10.1007/s44155-025-00207-3.

Agarwal, A., Mulgund, A., Hamada, A. and Chyatte, M.R. (2015). A unique view on male infertility around the globe. *Reproductive Biology and Endocrinology*, [online] 13(1). 10.1186/s12958-015-0032-1.

Arya, S.T. and Dibb, B. (2016). The experience of infertility treatment: the male perspective. Human Fertility, 19(4), pp.242–248. 10.1080/14647273.2016.1222083.

Barratt, C.L.R., Björndahl, L., De Jonge, C.J., Lamb, D.J., Osorio Martini, F., McLachlan, R., Oates, R.D., van der Poel, S., St John, B., Sigman, M., Sokol, R. and Tournaye, H. (2017). The diagnosis of male infertility: an analysis of the evidence to support the development of global WHO guidance—challenges and future research opportunities. *Human Reproduction Update*, [online] 23(6), pp.660–680. 10.1093/humupd/dmx021.

Biggs, S.N., Halliday, J. and Hammarberg, K. (2024). Psychological consequences of a diagnosis of infertility in men: a systematic analysis. Asian Journal of Andrology, 26(1), pp.10–19. 10.4103/aja202334.

Boivin, J., Oğuz, M., Duong, M., Cooper, O.R., Filipenko, D., Markert, M., Samuelsen, C. and Lenderking, W.R. (2023). Emotional reactions to infertility diagnosis: thematic and natural language processing analyses of the 1000 Dreams survey. Reproductive Biomedicine Online, 46(2), pp.399–409. 10.1016/j.rbmo.2022.08.107.

Brochu, F., Robins, S., Miner, S.A., Grunberg, P.H., Chan, P., Lo, K., Holzer, H.E.G., Mahutte, N., Ouhilal, S., Tulandi, T. and Zelkowitz, P. (2019). Searching the Internet for Infertility Information: A Survey of Patient Needs and Preferences. Journal of Medical Internet Research, 21(12), p.e15132. 10.2196/15132.

Burr, J.A. (2010). To name or not to name? An overview of the social and ethical issues raised by removing anonymity from sperm donors. Asian Journal of Andrology, 12(6), pp.801–806. 10.1038/aja.2010.60.

Cao, H., Liu, S., Cui, S., Nie, H., Liu, X. and Qin, W. (2025). From stress signals to fertility challenges: the role of damage-associated molecular patterns in male reproduction. Frontiers in Immunology, 16. 10.3389/fimmu.2025.1598451.

Carron-Arthur, B., Cunningham, J.A. and Griffiths, K.M. (2014). Describing the distribution of engagement in an Internet support group by post frequency: A comparison of the 90-9-1 Principle and Zipf’s Law. Internet Interventions, 1(4), pp.165–168. 10.1016/j.invent.2014.09.003.

Dooley, M., Nolan, A. and Sarma, K.M. (2011). The psychological impact of male factor infertility and fertility treatment on men: a qualitative study. The Irish Journal of Psychology, 32(1-2), pp.14–24. 10.1080/03033910.2011.611253.

Fisher, J.R. and Hammarberg, K. (2012). Psychological and social aspects of infertility in men: an overview of the evidence and implications for psychologically informed clinical care and future research. *Asian Journal of Andrology*, [online] 14(1), pp.121–129. 10.1038/aja.2011.72.

Furini, C., Costantino, F., Granata, A.R., Spaggiari, G., Santi, D. and Simoni, M. (2025). Still counting sperm? Why novel, truly informative measurements of testis function in male infertility are urgently needed. Endocrine, 90(3), pp.1067–1078. 10.1007/s12020-025-04453-y.

Grootendorst, M. (2022). BERTopic: Neural topic modeling with a class-based TF-IDF procedure. *arXiv*. 10.48550/arxiv.2203.05794.

Hanna, E. and Gough, B. (2016a). Emoting infertility online: A qualitative analysis of men’s forum posts. *Health: An Interdisciplinary Journal for the Social Study of Health*, Illness and Medicine, 20(4), pp.363–382. 10.1177/1363459316649765.

Hanna, E. and Gough, B. (2016b). Searching for help online: An analysis of peer-to-peer posts on a male-only infertility forum. Journal of Health Psychology, 23(7), pp.917–928. 10.1177/1359105316644038.

Hanna, E., Gough, B. and Hudson, N. (2018). Fit to father? Online accounts of lifestyle changes and help-seeking on a male infertility board. *Sociology of Health & Illness*, [online] 40(6), pp.937–953. 10.1111/1467-9566.12733.

Hartmann, J. (2022). *Emotion english DistilRoBERTa-base*. [online] *Hugging Face*. Available at: https://huggingface.co/j-hartmann/emotion-english-distilroberta-base/ [Accessed 8 Nov. 2025].

Heitmann, A. (2025). GitHub - ArthurHeitmann/arctic_shift: Making Reddit data accessible to researchers, moderators and everyone else. Interact with the data through large dumps, an API or web interface. [online] GitHub. Available at: https://github.com/ArthurHeitmann/arctic_shift [Accessed 11 May 2025].

Hirschberg, J. and Manning, C.D. (2015). Advances in natural language processing. *Science*, [online] 349(6245), pp.261–266. 10.1126/science.aaa8685.

Holley, S.R., Pasch, L.A., Bleil, M.E., Gregorich, S., Katz, P.K. and Adler, N.E. (2015). Prevalence and predictors of major depressive disorder for fertility treatment patients and their partners. Fertility and Sterility, 103(5), pp.1332–1339. 10.1016/j.fertnstert.2015.02.018.

Houston, B.J., Riera-Escamilla, A., Wyrwoll, M.J., Salas-Huetos, A., Xavier, M.J., Nagirnaja, L., Friedrich, C., Conrad, D.F., Aston, K.I., Krausz, C., Tüttelmann, F., O’Bryan, M.K., Veltman, J.A. and Oud, M.S. (2022). A systematic review of the validated monogenic causes of human male infertility: 2020 update and a discussion of emerging gene-disease relationships. *Human Reproduction Update*, [online] 28(1), pp.15–29. 10.1093/humupd/dmab030.

Huang, B., Wang, Z., Kong, Y., Jin, M. and Ma, L. (2023). Global, regional and national burden of male infertility in 204 countries and territories between 1990 and 2019: an analysis of global burden of disease study. *BMC Public Health*, [online] 23(1). 10.1186/s12889-023-16793-3.

Kalampalikis, N., Doumergue, M. and Zadeh, S. (2018). Sperm donor regulation and disclosure intentions: Results from a nationwide multi-centre study in France. Reproductive Biomedicine & Society Online, 5, pp.38–45. 10.1016/j.rbms.2018.02.001.

Köckritz, J., İlgen, B., Cohrdes, C. and Hattab, G. (2025). Current applications and future directions in natural language processing for news media and mental health. Scientific Reports, 15(1). 10.1038/s41598-025-18413-z.

Krausz, C. and Riera-Escamilla, A. (2018). Genetics of male infertility. *Nature Reviews Urology*, [online] 15(6), pp.369–384. 10.1038/s41585-018-0003-3.

Kuroda, S., Usui, K., Sanjo, H., Takeshima, T., Kawahara, T., Uemura, H. and Yumura, Y. (2020). Genetic disorders and male infertility. *Reproductive Medicine and Biology*, [online] 19(4), pp.314–322. 10.1002/rmb2.12336.

Li, J., Li, K., Zhang, M., Chen, J., Chen, M., Dong, W., Yu, W., Lei, L., Huang, Y., Yang, H., Ren, P., Zou, Q. and Deng, L. (2025). Male Infertility Management: A Critical Appraisal of Clinical Practice Guidelines With the AGREE II Instrument. American Journal of Men’s Health, 19(5), p.15579883251380203–15579883251380203. 10.1177/15579883251380203.

Lin, J.W. and Shorey, S. (2023). Online peer support communities in the infertility journey: A systematic mixed-studies review. International Journal of Nursing Studies, 140, p.104454. 10.1016/j.ijnurstu.2023.104454.

Mitrakas, A.G., Alexiadi, C.-A., Gargani, S., Alexiadis, T., Alexopoulou, S.-P., Pagonopoulou, O. and Lambropoulou, M. (2025). Chromosomal Roadblocks in Male Fertility: Mechanisms, Risk Factors and Syndromes. *Medicina*, [online] 61(10), p.1864. 10.3390/medicina61101864.

Morgan, J., Trigo, A. and Davies, K. (2020). Assessing the change in infertility patients’ social media use during COVID-19 related clinic closures. Fertility and Sterility, 114(3), p.e545. 10.1016/j.fertnstert.2020.09.084.

Mueller, A. (2020). word_cloud. [online] GitHub. Available at: https://github.com/amueller/word_cloud [Accessed 11 Oct. 2025].

O’Connell, S.B.L., Gelgoot, E.N., Grunberg, P.H., Schinazi, J., Da Costa, D., Dennis, C.-L., Rosberger, Z. and Zelkowitz, P. (2021). ‘I felt less alone knowing I could contribute to the forum’: psychological distress and use of an online infertility peer support forum. Health Psychology and Behavioral Medicine, 9(1), pp.128–148. 10.1080/21642850.2021.1884556.

Olesen, I.A., Andersson, A.-M., Aksglaede, L., Skakkebaek, N.E., Rajpert–de Meyts, E., Joergensen, N. and Juul, A. (2017). Clinical, genetic, biochemical, and testicular biopsy findings among 1,213 men evaluated for infertility. Fertility and Sterility, 107(1), pp.74–82.e7. 10.1016/j.fertnstert.2016.09.015.

Osadchiy, V., Jiang, T., Mills, J.N. and Eleswarapu, S. (2020). Low Testosterone on Social Media: Application of Natural Language Processing to Understand Patients’ Perceptions of Hypogonadism and Its Treatment. Journal of Medical Internet Research, 22(10), pp.e21383–e21383. 10.2196/21383.

Osadchiy, V., Mills, J.N. and Eleswarapu, S.V. (2020). Understanding Patient Anxieties in the Social Media Era: Qualitative Analysis and Natural Language Processing of an Online Male Infertility Community. Journal of Medical Internet Research, 22(3), p.e16728. 10.2196/16728.

Pallotti, F., Barbonetti, A., Rastrelli, G., Santi, D., Corona, G. and Lombardo, F. (2022). The impact of male factors and their correct and early diagnosis in the infertile couple’s pathway: 2021 perspectives. Journal of Endocrinological Investigation. 10.1007/s40618-022-01778-7.

Patel, D., Blandford, A., Warner, M., Shawe, J. and Stephenson, J. (2019). ‘I feel like only half a man’. Proceedings of the ACM on Human-Computer Interaction, 3(CSCW), pp.1–20. 10.1145/3359184.

Patel, N., Sharma, R., Lingasamy, P., Sundararajan, V., S, S.L. and Modhukur, V. (2026a). Temporal evolution of digital health communication in Rheumatoid Arthritis: A longitudinal NLP analysis of reddit discussions (2018–2024). PLOS One, 21(1), p.e0341006. 10.1371/journal.pone.0341006.

Patel, N., Sharma, R., Prakash Lingasamy, Sundararajan, V., Sajitha Lulu Sudhakaran and Vijayachitra Modhukur (2026b). Understanding user perceptions of DeepSeek: insights from sentiment, topic and network analysis using a Reddit-based study. Frontiers in Artificial Intelligence, 8. 10.3389/frai.2025.1703949.

Pearson, K. (1900). On the Criterion That a Given System of Deviations from the Probable in the Case of a Correlated System of Variables Is Such That It Can Be Reasonably Supposed to Have Arisen from Random Sampling. *The London*, Edinburgh, and Dublin Philosophical Magazine and Journal of Science, 50(302), pp.157–175. 10.1080/14786440009463897.

Pozzi, E., Corsini, C. and Salonia, A. (2024). Medical therapy for male infertility. Current Opinion in Urology. 10.1097/mou.0000000000001231.

Sahoo, S., Das, A., Dash, R., Behera, A., Mishra, N. and Bal, K. (2025). The Psychological Impact of Male Infertility: A Narrative Review. Cureus. 10.7759/cureus.89453.

Sax, M.R. and Lawson, A.K. (2022). Emotional Support for Infertility Patients: Integrating Mental Health Professionals in the Fertility Care Team. Women, 2(1), pp.68–75. 10.3390/women2010008.

Schlegel, P.N., Sigman, M., Collura, B., De Jonge, C.J., Eisenberg, M.L., Lamb, D.J., Mulhall, J.P., Niederberger, C., Sandlow, J.I., Sokol, R.Z., Spandorfer, S.D., Tanrikut, C., Treadwell, J.R., Oristaglio, J.T. and Zini, A. (2021a). Diagnosis and treatment of infertility in men: AUA/ASRM guideline part I. Fertility and Sterility, 115(1), pp.54–61. 10.1016/j.fertnstert.2020.11.015.

Schlegel, P.N., Sigman, M., Collura, B., De Jonge, C.J., Eisenberg, M.L., Lamb, D.J., Mulhall, J.P., Niederberger, C., Sandlow, J.I., Sokol, R.Z., Spandorfer, S.D., Tanrikut, C., Treadwell, J.R., Oristaglio, J.T. and Zini, A. (2021b). Diagnosis and Treatment of Infertility in Men: AUA/ASRM Guideline PART II. Journal of Urology, 205(1), pp.44–51. 10.1097/ju.0000000000001520.

Sellke, N., Jesse, E., Callegari, M., Muncey, W., Harris, D., Edwins, R., Pominville, R., Ghayda, Ramy Abou, Loeb, A. and Thirumavalavan, N. (2023). Is Reddit a reliable source for information on erectile dysfunction treatment? *International Journal of Impotence Research*, [online] 35(5), pp.484–489. 10.1038/s41443-022-00586-0.

Simbar, M., Rashidi, F., Taherian, R., Ghasemi, V., Kalhor, M. and Kiani, Z. (2025). Is stress different in infertile women and men? A systematic review and meta-analysis. BMC Public Health, 25(1). 10.1186/s12889-025-24964-7.

Sollender, G.E., Jiang, T., Finkelshtein, I., Osadchiy, V., Zheng, M.H., Mills, J.N., Singer, J.S. and Eleswarapu, S.V. (2024). Understanding Pediatric Experiences With Symptomatic Varicoceles: Mixed Methods Study of an Online Varicocele Community. *JMIR Formative Research*, [online] 8, p.e50141. 10.2196/50141.

Treadgold, B.M., Coulson, N.S., Campbell, J.L., Lambert, J. and Pitchforth, E. (2025). Quality and misinformation about health conditions in peer online support groups: Scoping review of information and advice (Preprint). Journal of Medical Internet Research. 10.2196/71140.

Wang, K., Xu, Y., Qin, N., Guo, Y., Bai, J. and Li, Z. (2025). Effect of Donor Sperm on Quality of Life in Patients with Severe Oligoasthenosperm After ICSI Failure. *Journal of Multidisciplinary Healthcare*, Volume 18, pp.123–132. 10.2147/jmdh.s494319.

Zadeh, S., Freeman, T. and Golombok, S. (2016). Absence or presence? Complexities in the donor narratives of single mothers using sperm donation. Human Reproduction, 31(1), pp.117–124. 10.1093/humrep/dev275.

Zafar, M.I., Mills, K.E., Baird, C.D., Jiang, H. and Li, H. (2023). Effectiveness of Nutritional Therapies in Male Factor Infertility Treatment: A Systematic Review and Network Meta-analysis. Drugs. [online] 10.1007/s40265-023-01853-0.

